# Assessing the Effects of Behavioral Circadian Rhythm Disruption in Shift-Working Police Academy Trainees

**DOI:** 10.1101/2021.07.23.21261052

**Authors:** Melissa L. Erickson, Will Wang, Julie Counts, Leanne M. Redman, Daniel Parker, Janet L. Huebner, Jessilyn Dunn, William E. Kraus

## Abstract

Night shift work, characterized by behavioral circadian disruption, increases cardiometabolic disease risk. Our long-term goal is to develop a novel methodology to quantify behavioral circadian disruption in field-based settings and to explore relations to four metabolic salivary biomarkers of circadian rhythm. This pilot study enrolled 36 police academy trainees to test the feasibility of using wearable activity trackers to assess changes in behavioral patterns. Using a two-group observational study design, participants completed in-class training during dayshift for six weeks followed by either dayshift or nightshift field-training for six weeks. We developed a novel data-post processing step that improves sleep detection accuracy of sleep episodes that occur during daytime. We next assessed changes to resting heart rate (RHR) and sleep regularity index (SRI) during dayshift versus nightshift field training. Secondarily, we examined changes in field-based assessments of salivary cortisol, uric acid, testosterone, and melatonin during dayshift versus nightshift. Compared to dayshift, nightshift workers experienced larger changes to resting heart rate, sleep regularity index (indicating reduced sleep regularity), and alternations to sleep/wake activity patterns accompanied by blunted salivary cortisol. Salivary uric acid, testosterone, and melatonin did not change. These findings show that nightshift work—a form of behavioral circadian rhythm disruption—was detectable in police trainees using activity trackers alone and in combination with a specialized data analysis methodology.

**KEY POINTS:**

- Night shift work increases cardiometabolic disease risk and this may be a consequence of behavioral circadian misalignment.
- To advance this hypothesis, methodologies to quantify behavioral irregularities during nightshift in field-based settings are needed.
- In this pilot study, commercially available activity trackers combined with a novel data processing step were used to assess alterations in sleep/wake patterns in police trainees during dayshift versus nightshift.
- We also explored relations with four metabolic salivary biomarkers of circadian rhythm during dayshift versus nightshift.
- Compared to dayshift, nightshift resulted in larger perturbations of resting heart rate, sleep regularity index (indicating reduced regularity), and alterations in sleep and activity patterns; this was accompanied by blunted cortisol.
- This novel data processing step extends commercially available technology for successful application in real-world shift work settings.

## INTRODUCTION

Diverse occupational sectors—transportation, healthcare, manufacturing, and public safety—rely on shiftwork schedules in order to meet work sector demands. Mounting evidence suggests circadian disruptions caused by shiftwork schedules result in increased chronic disease risk (Antunes *et al*., 2010; Pan *et al*., 2011; Lieu *et al*., 2012; Barbadoro *et al*., 2013; Depner *et al*., 2014; Vetter *et al*., 2016; Manohar *et al*., 2017; Shan *et al*., 2018; Gao *et al*., 2019; Dutheil *et al*., 2020; Rivera *et al*., 2020; Schilperoort *et al*., 2020; Maidstone *et al*., 2021). For example, shiftwork is associated with obesity, type 2 diabetes (Antunes *et al*., 2010; Shan *et al*., 2018; Gao *et al*., 2019), hypertension (Manohar *et al*., 2017), dyslipidaemia (Dutheil *et al*., 2020), asthma (Maidstone *et al*., 2021), as well as increased breast cancer risk and stroke (Rivera *et al*., 2020). While the relationship between shiftwork and chronic disease susceptibility is likely complex, it is hypothesized that temporal misalignment between the internal circadian clock and worktimes play a role.

To advance our understanding of the relationship between circadian disruption introduced by shiftwork and increased chronic disease risk, a feasible, straightforward methodology for assessing field-based behavioral circadian disruption is needed. This requisite was recently highlighted in a white paper summarizing discussions at the 2018 Sleep Research Society’s sponsored workshop, “International Biomarkers Workshop and Wearables in Sleep and Circadian Science” (Depner *et al*., 2020). The widespread development of commercially available activity trackers affords researchers new opportunities to survey novel behavioral patterns in community settings that can be linked to key health indicators (Shcherbina *et al*., 2017). Wrist-worn smart watches provide information on behavioral regularity of when an individual sleeps and exercises.

Current activity tracker technology is optimized for use in settings when “typical” sleep/wake behaviors occur, in that devices are more likely to accurately detect activity during daytime hours and sleep during nighttime hours. However, this may be problematic used in the shift work setting. Shiftwork requires an individual to be active during the nighttime hours and sleep during daytime hours. These misaligned behaviors are likely to go undetected, leading to inaccurate quantification. This shortcoming may be overcome by developing a novel data post-processing step that removes external clock time bias, thereby increasing sleep label detection accuracy in the shiftwork setting.

We anticipated that proprietary sleep algorithms originally developed for use by consumers with regular sleep patterns might perform poorly during night shiftwork: daytime sleep episodes would go undetected. Therefore, the first aim of this study was to develop a novel algorithm for sleep detection that is not biased by external clock time, in a sample of shift working police trainees. The second aim was to assess the feasibility of concurrent field-based salivary sampling to detect changes in known biomarkers of circadian patterns. We hypothesized that our novel algorithm would accurately detect daytime sleep episodes that are missed by commercial technology; and, secondly, that nightshift work would be reflected by aberrations in biological samples (cortisol, uric acid, testosterone, and melatonin).

## METHODS

### Study Design

This was a two-group observational, repeated measures study design, leveraging the established schedule followed by 36 police recruits. Schedules of police recruits involve 24 weeks of in-class training followed by 14 weeks of field-training. This pilot study lasted approximately twelve weeks and occurred during the last six weeks of in-class training (baseline phase) and the first six weeks of field-training. During in-class training, classes were held Monday through Friday during daytime (7:30 AM-5:00 PM) hours; this represented normal circadian alignment. This baseline phase was subsequently followed by six weeks of field training. During the field-training phase, 13 participants maintained a normal daytime schedule, representing circadian alignment and 14 participants switched to night shift work, representing circadian misalignment (Figure 1). During the second phase (circadian misalignment), trainees were assigned to one of the following four shift work schedules:

**Figure 1:**
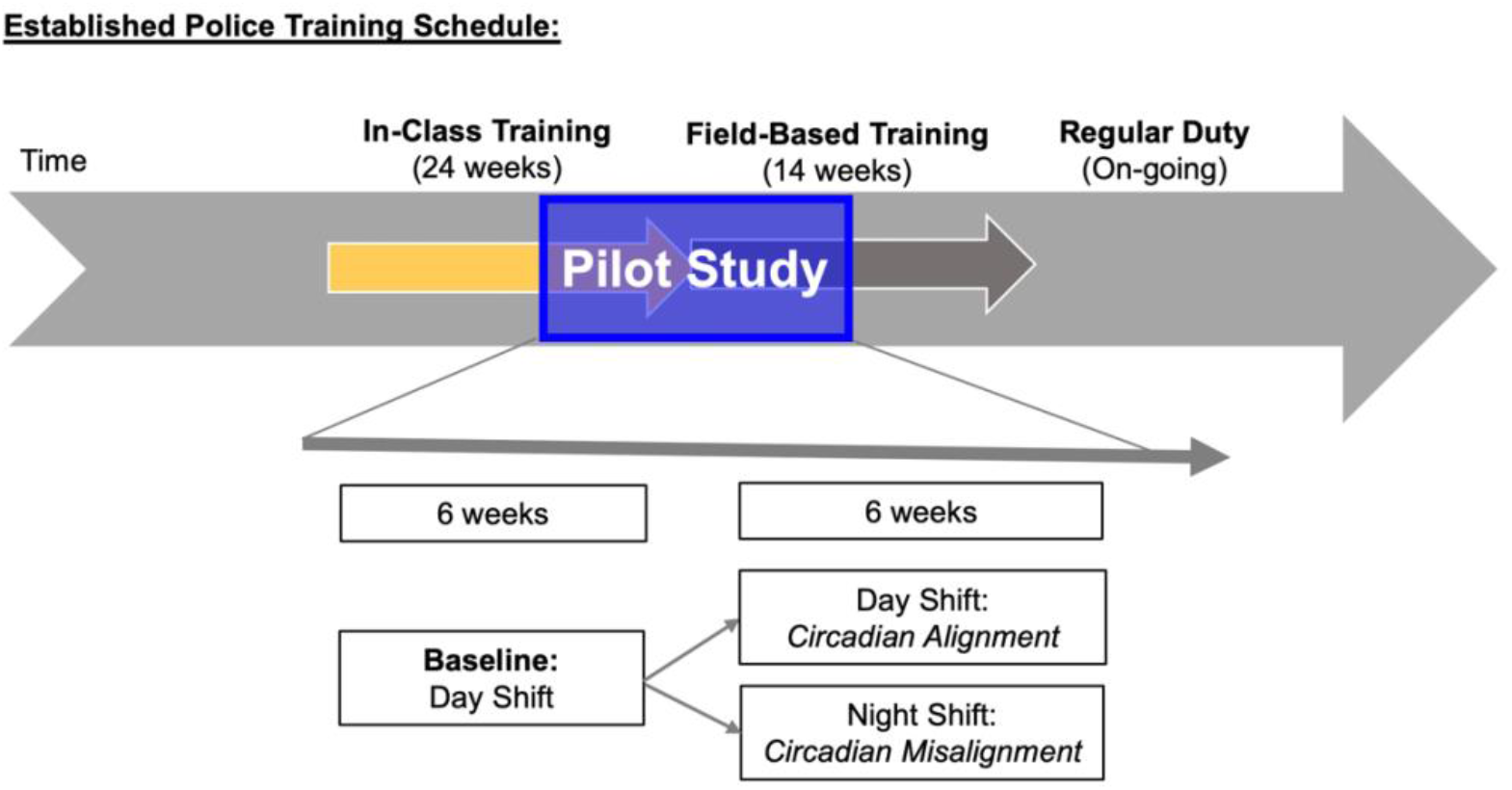
Study Design.

*Schedule A: 6 AM-5 PM (circadian alignment)*

*Schedule B: 10 AM-9 PM (circadian alignment)*

*Schedule C: 4 PM-3 AM (circadian misalignment)*

*Schedule D: 8 PM-7 AM (circadian misalignment)*

Two of these four field-training schedules (A and B) align with the 24h day/night cycle and represented a maintenance of behavioral circadian alignment. One participant engaged in office work continued to follow a 8 AM-5 PM schedule. The other two schedules (C and D) were misaligned with the day/night cycle and represented acute circadian misalignment. Work schedules were maintained for four consecutive days, followed by four consecutive days off. Activity monitors were worn continuously, and thus capture behavior during both the in-class training phase and the field training phase. Three salivary samples were collected during the in-class training phase and six samples were collected during the field training phase, totaling 9 salivary samples for each participant. This study design was advantageous because it controls for the job transition from in-class training to field-training due to nightshift and dayshift transition comparisons.

### Ethical Approval

This study was approved by the Duke University Health System Institutional Review Board for Clinical Investigations (IRB# Pro00077319). All participants provided written informed consent prior to study participation.

### Participants

Study inclusion criteria were as follows: 1) enrolled in a local public safety training program and 2) owned a smartphone. We conducted on-site recruitment events to raise general study awareness by partnered with a local policy department. We presented the study to a total of 77 trainees, or four academy classes and enrolled 36 participants. Participants provided informed consent electronically using a secure web application (REDcap).

### Study Protocol

As a field-based study, all assessments were collected outside of the laboratory. After providing informed consent, participants were instructed on use of the activity tracker (Garmin vívosmart® HR, Olathe, KS) and supplied with six self-administered saliva collection kits, using either drool sampling (SalivaBio Passive Drool, Salimetrics^®^) or oral swab method (SalivaBio Oral Swab, Salimetrics^®^), and then instructed on their use and the collection protocol.

### Activity Tracker Assessments

Activity monitors were worn on the wrist 24/7 (except for when the watch was being charged). The Garmin vívosmart® HR recorded observations of activity level, heart rate, and algorithmically-generated sleep/wake labels every 15 minutes, totaling 96 measurements per person during a 24h period. Wear time was required to be at least 80% over a given 24h period and individuals meeting this criterion for at least 50% of the days were considered complete and included in the analysis. From these data, changes in resting heart rate and in sleep regularity using the methods described subsequently were evaluated.

#### Novel Sleep Labelling Method Development

Garmin vívosmart® HR relies on user input of anticipated regular bedtime—a key input to the sleep detection algorithm. However, shift workers followed irregular sleep/wake patterns and this may potentially contribute to inaccuracies in sleep detection, particularly during daytime hours. We posit that a novel sleep labeling method that does not require user input information, such as anticipated bedtime, will increase sleep detection accuracy in the shift work setting. Thus, we developed a novel logistic regression-based sleep labeling algorithm that relies on heart rate and activity data—rather than anticipated bedtime—as input information to detect sleep episodes. Specifically, we developed a model that labels 15-minute epochs as sleep or wake based on new input information—heart rate and activity data—collected by Garmin vívosmart® HR and application.

To accomplish this, we first defined our ground truth data set using the following rationale. We acquired reliable sleep periods detected by the Garmin vívosmart® HR and application. Given that Garmin technology is optimized for a typical circadian aligned schedule, we assumed that Garmin vívosmart® HR sleep labels (positive labels) collected during in-class training (which follows a daytime schedule) were reliable. Specifically, reliable wake labels were defined as periods 4 to 8 hours before the sleep period start and 4 to 8 hours after the sleep period end. Next, the ground truth dataset was split into training (n=148256) and test sets (n=37064) for algorithm development.

#### Resting Heart Rate

Daily resting heart rate was calculated as the mean heart rate at rest, or when the maximum Motion Intensity < 3. Motion Intensity was derived from minute-level accelerometry data and is an aggregate measure of overall activity level for each 15-minute epoch. Motion Intensity takes integer values between 0 and 7 inclusively, with 0 corresponding to stillness and higher scores corresponding to more activity.

#### Sleep Regularity Index

We calculated a sleep regularity index, using sleep/wake labels obtained after the sleep labelling algorithm, to quantify day-to-day sleep regularity over the course of five consecutive days. This a previously established index that ranges from 0-100, in which a greater value indicated increased sleep regularity. The equation for calculation of the sleep regularity index has been described previously. It was used initially used on ActiGraph’s sleep/wake label data streams; and therefore, could be easily applies to sleep/wake labels derived from Garmin vívosmart® HR activity and heart rate data (Lunsford-Avery *et al*., 2018) for use in the current study.

### Salivary Assessments

Saliva samples were self-collected using either the cheek cotton swab method or the passive drool method, in which saliva is passed into a collection container via straw. Participants stored saliva samples in their home -20°C freezer until collected by study staff at the following protocol visit. Samples were then stored at -80°C until batched analyses. During in-class training, which represents baseline, participants collected three samples: before bed, upon waking, and 30 minutes after waking (sample must be collected within 60 minutes after waking to be included in final analysis) on a workday (totaling 3 samples). During field-training, participants collected three samples at the same behavioral events on a workday and non-workday (totaling 6 samples). The workday and non-workday samples were averaged, to represent the behavioral timepoints for field training.

The differences between in-class training and field-training (average of workday and non-workday) were calculated for each behavioral time point:

a. before bed_(in class-training)_ – before bed_(field training; average of workday and non-workday)_
b. upon waking _(in class-training)_ – upon waking_(field training; average of workday and non-workday)_
c. wake + 30 min_(in class-training)_ – wake +30 min_(field training; average of workday and non-workday)_

Assessment timepoints are shown in Figure 2. These calculations were performed on salivary biomarkers: cortisol, uric acid, testosterone, and melatonin. Next, we compared to deltas between in-class dayshift versus those during in-class nightshift.

**Figure 2:**
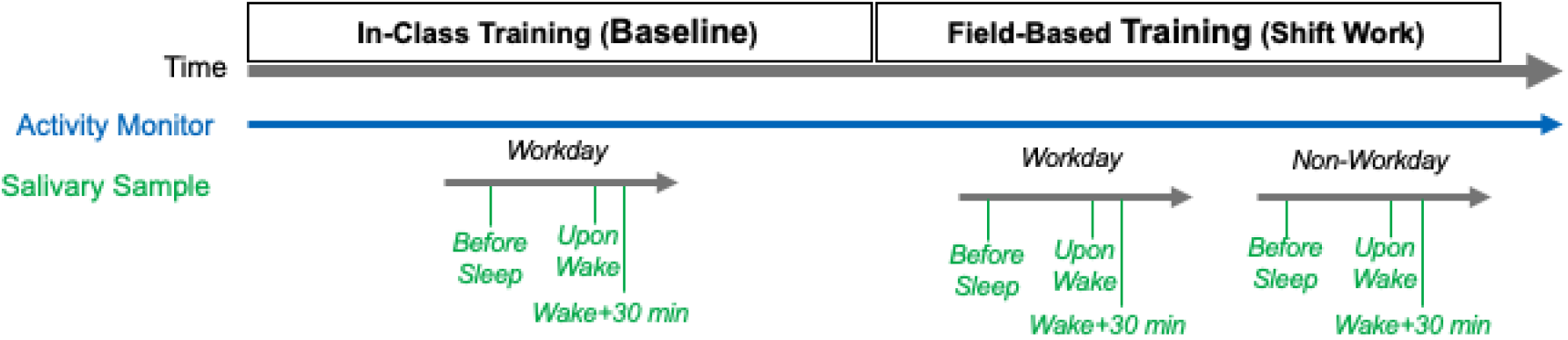
Activity Monitor and Salivary Sample Assessment Timeline.

#### Salivary Circadian Biomarker Assays

To assess salivary biomarkers, manufacturer’s instructions were followed using commercially available immunoassay kits; salivary cortisol (Salimetrics #1-3002), salivary uric acid (Salimetrics #1-3802), salivary testosterone (Salimetrics #1-2402), and salivary melatonin (Salimetrics #1-3402). To minimize batch effects, all three behavioral timepoints from a participant were analyzed on the same plate (e.g., saliva sample collected at baseline, upon waking, and wake + 30 min during both in-class training and field-based training). Manufacturer-provided controls were run in duplicate on each plate in order to assess intra- and inter-assay variability and to establish an acceptable control range. Lab personnel were blinded to study condition.

### Statistical Analysis

Data are presented as mean ± standard deviation unless otherwise noted. Python 3.6 (packages statmodels 0.11.0 and pingouin 0.3.11) was used for statistical analyses.

#### Aim 1

Novel Algorithm Performance: We tested agreement between our novel algorithm and reliable Garmin sleep labels by evaluating the following: testing and training accuracy, testing F1-score, and testing ROC-AUC. To accomplish this, we used four performance evaluation models: logistic regression, random forest, adaboost, and support vector machine (radial basis function). We considered 0.90 testing F1-score and testing ROC-AUC as acceptable performance.

#### Aim 2

Within each group, differences between in-class training versus field-training were determined using the Wilcoxon signed-rank test for activity monitor measures (resting heart rate and sleep regularity index) and salivary measures (cortisol, uric acid, testosterone, and melatonin). Between group differences (circadian misalignment vs. circadian alignment) during the transition from in-class to field-training were determined using the non-parametric Kruskal Wallis test for both activity monitor measures (resting heart rate and sleep regularity index) and salivary measures (cortisol, uric acid, testosterone, and melatonin). The significance threshold was *P*<0.05. Salivary biomarkers were adjusted for multiple comparisons using Bonferroni corrections. Outliers were identified by Grubb’s test.

## RESULTS

### Participant Characteristics

The study cohort was predominately male (67%; 18 M/9 F). The mean age was 28 years old (6.2) ranging from 21 to 47 yrs. The mean BMI was 27 kg/m^2^ (± 3.4) ranging from 21 to 33 kg/m^2^. A consort figure is shown in Figure 3. Nine of 36 enrolled participants did not have data due to various reasons (e.g. lost to follow up, did not follow sample collection instructions; Fig 3).

**Figure 3:**
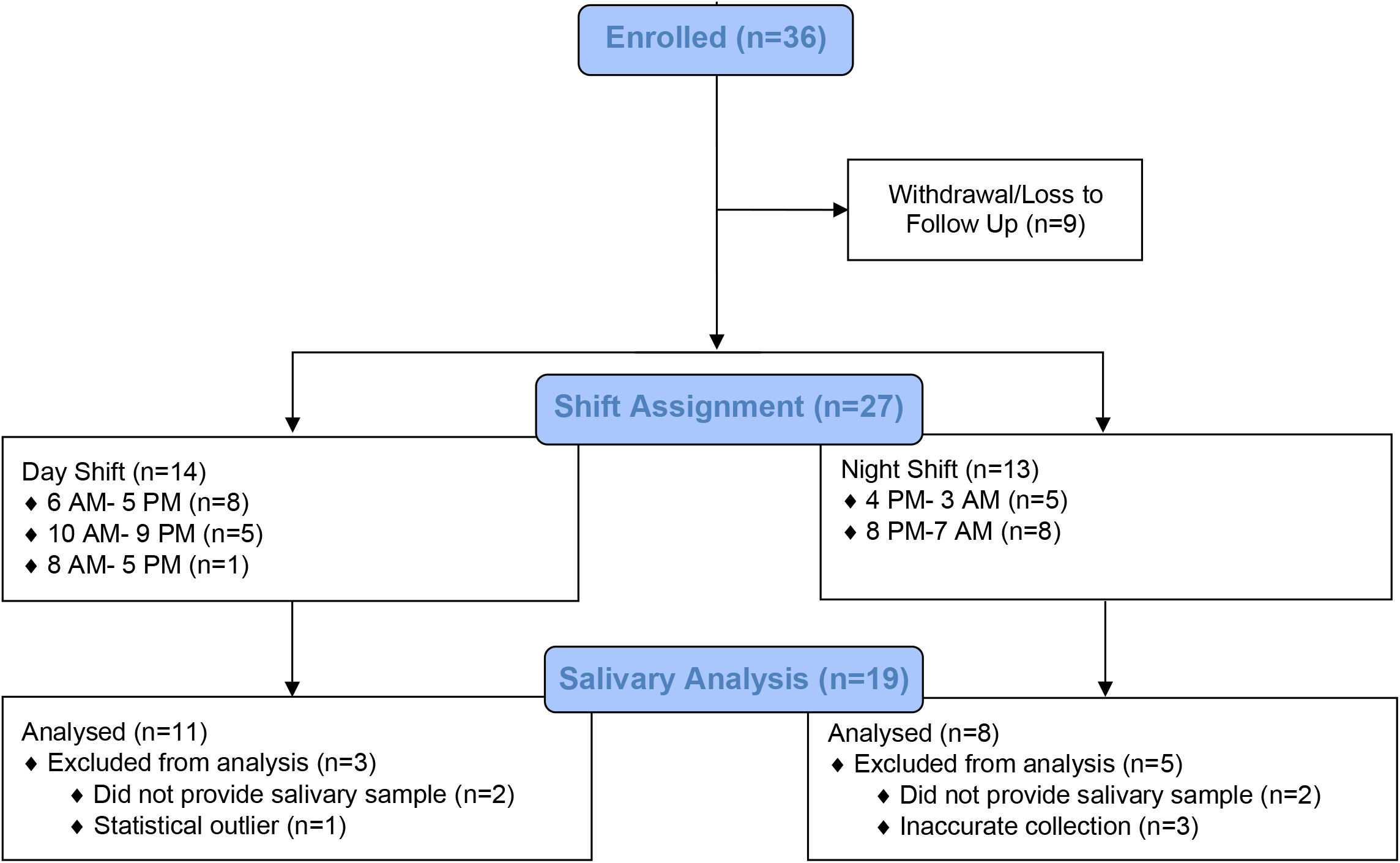
Consort Diagram.

### Activity Tracker Sleep Detection Performance During Shift Work

Of the 27 participants that completed both phases, we had complete data for 25 participants. We excluded activity tracker data from two participants as they did not meet our wear time criteria.

To determine whether we can rely on sleep provided by Garmin, we evaluated the performance of Garmin sleep detection during both dayshift and nightshift. We expect trainees to have at least one main sleep event every 24h, which we defined as the largest block of time spent asleep or in bed exceeding 4h, regardless of circadian alignment. We compared the number of Garmin-generated sleep periods to this expectation and observed that during the day shift field-training (circadian alignment), 89.7% of main sleep events were detected. However, during the night shift field-training (circadian misalignment) only 49.7% of main sleep events were detected. We interpreted this to mean activity tracker proprietary algorithms have high sleep detection accuracy used during typical circadian aligned schedule, but poor sleep detection performance during circadian misalignment. These findings re-affirm the need to improve algorithm sleep detection performance during nightshift.

### Novel Sleep Labeling Development

Our algorithm demonstrated high epoch-by-epoch prediction accuracy on the test dataset, with logistic regression achieving a testing accuracy, or level of agreement with the Garmin algorithm, of 94% (Table 1). While all four models demonstrated high level of agreement, we ultimately chose logistic regression because of model simplicity and less risk of overfitting. We then used the logistic regression model to determine sleep versus wake labels for each epoch during nightshift work (circadian misalignment) and imputed the labels previously missed by the activity tracker’s proprietary sleep detection algorithm.

**Table 1:** Novel Sleep Labelling Method Performance Statistics.

|  | Training Accuracy | Testing Accuracy | Testing F1-Score | Testing ROC-AUC |
| --- | --- | --- | --- | --- |
| Logistic Regression (CV) | **0.94** | **0.94** | **0.94** | **0.98** |
| Random Forest | 0.95 | 0.95 | 0.95 | 0.98 |
| Adaboost | 0.94 | 0.94 | 0.94 | 0.98 |
| SVM (rbf) | 0.94 | 0.94 | 0.95 | 0.95 |

SVM (rbf): Support vector machine (radial basis function).

Figure 4 compares sleep labeling detected by activity tracker propriety software (gray shading) versus sleep labeling detected by our novel sleep labeling method (overlaid orange shading) over the course of four days, plotted as heart rate (Panel A), Mean Motion Intensity (Panel B) and Max Motion Intensity (Panel C). These data shown that activity tracker labeling detected a main sleep event during the first 24h period but missed sleep events during subsequent three nights. In contrast, our sleep labeling method detects a main sleep event for each 24h period. These four days were chosen arbitrarily.

**Figure 4:**
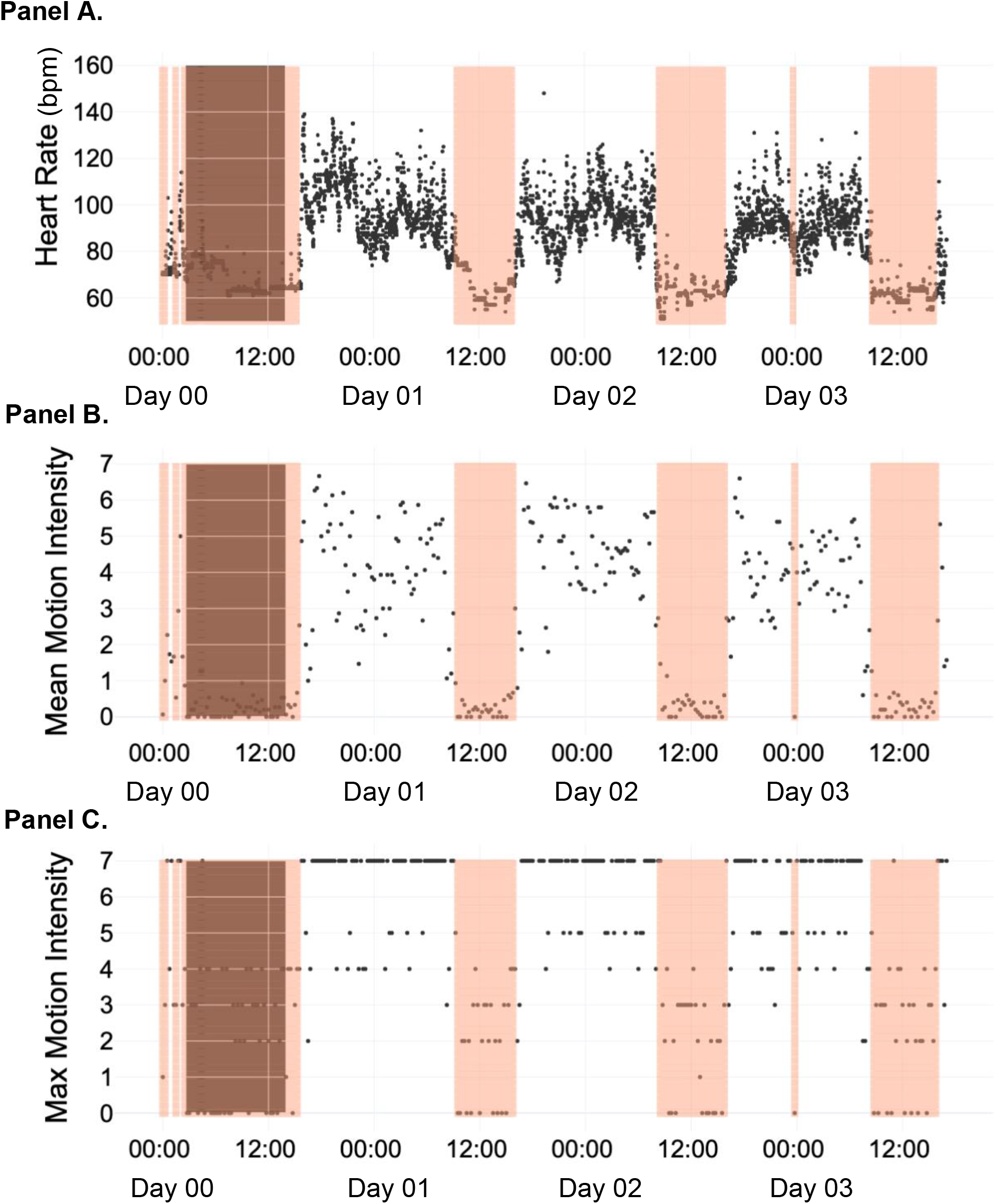
Comparison of Activity Tracker Sleep Labeling Vs. Novel Sleep Labeling Method. Heart rate, mean motion intensity and max motion intensity derived from activity monitor collected over a typical span of four days for an individual police trainee during field-training. X-axis displays time (12h increments indicated). Data points are shown at a frequency of every 15 minutes. **Panel A** shows heart rate (beats per minute) over the course of four days represented by black symbols. **Panel B** shows mean motion intensity over the course of four days represented by black symbols. **Panel C** shows max motion intensity over the course of four days represented by black symbols. The gray shaded area is the Garmin-detected sleep period. The orange shaded area denotes sleep period detected by our novel sleep labeling method.

### Activity Tracker Assessments

To determine whether physical activity and algorithm-derived sleep patterns across phases of the day were indicative of the occurrence of a circadian misalignment, we developed a polar plot to visualize activity and sleep behavior fluctuations during both daytime training versus night shift field-training, totaling 42 days. As shown (Figure 5), the polar plots depict behavioral pattern shifts relative to the external clock time over long durations (e.g., several weeks) and demonstrate the dramatic shift in the sleep/wake routine relative to the external clock time that is absent during dayshift work (Figure 5A) but present during night shiftwork (Figure 5B). Hence, we concluded that the following a nightshift schedule resulted in behavioral circadian misalignment.

**Figure 5:**
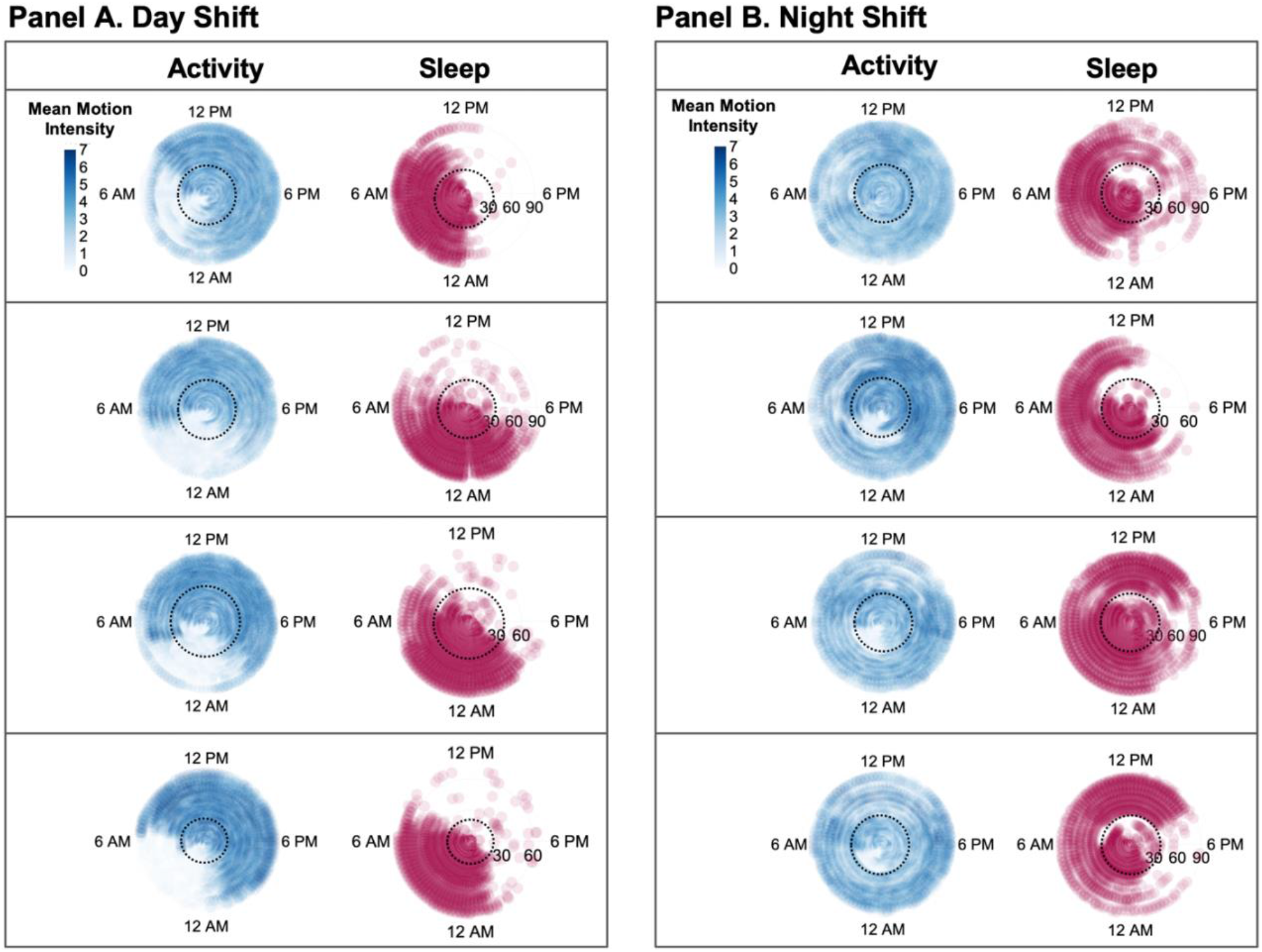
Polar plot display of activity patterns assessed using Garmin vívosmart® HR (shown in blue) and sleep patterns derived from the novel sleep labelling method (shown in red). Days are plotted on the radial axis for two consecutive observational periods (in-class training followed by field-training). Each activity and sleep data pair represent one participant. **Panel A** shows paired activity and sleep data from 4 participants assessed during dayshift circadian alignment. **Panel B** shows paired activity and sleep data from 4 participants during nightshift circadian misalignment. Black dashed circle indicates approximate timing of the transition from in-class training to field training. Intensity of the activity data are represented by increased pixel color intensity as indicated in the figure legend on the top level of both panels.

#### Resting Heart Rate

For the trainees remaining in circadian alignment, resting heart rate was 63.5 ± 6.4 bpm (beats per minute) during in-class training, and increased to 65.4 ± 6.7 bpm during field-training. Whereas for the trainees who underwent circadian misalignment, resting heart rate increased from 66.1 ± 4.5 during in-class training to 72.5 ± 6.0 bpm during field-training. Circadian misalignment resulted in a significantly larger change in resting heart rate (*P*=0.009, Table 2).

**Table 2.** Behavioral and Biological Changes During Shift Work.

| **Outcome Measures**<br>*Delta from In-Class Training to Field-Training* | **Dayshift:**<br>***Circadian Alignment*** | **Nightshift:**<br>***Circadian Misalignment*** | ***P Value*** |
| --- | --- | --- | --- |
| **Activity Tracker Assessments** | -- | -- | -- |
| Resting Heart Rate, bpm | 1.84 ± 2.33 | 6.48 ± 4.28 | 0.00898* |
| Sleep Regularity Index | 1.44 ± 10.8 | 9.49 ± 10.9 | 0.0500 |
| **Salivary Biomarker** |  |  | -- |
| Cortisol |  |  |  |
| before bed | -0.0018 ± 0.13 | -0.046 ± 0.11 | 0.142 |
| upon waking | -0.00056 ± 0.15 | -0.11 ± 0.21 | 0.310 |
| wake+30 min | 0.04 ± 0.18 | -0.32 ± 0.23 | 0.00156* |
| Uric Acid |  |  |  |
| before bed | 0.012 ± 1.32 | -0.29 ± 3.1 | 0.280 |
| upon wake | -1.01 ± 3.79 | 2.81 ± 3.47 | 0.128 |
| wake+30 min | -0.12 ± 1.85 | -0.52 ± 3.4 | 0.939 |
| Testosterone |  |  |  |
| before bed | -15.4 ± 70.0 | -27.2 ± 145 | 0.440 |
| upon wake | 232 ± 721 | 40.3 ± 83.5 | 0.866 |
| wake+30 min | -40.9 ± 126 | -3.32 ± 61.3 | 0.537 |
| Melatonin |  |  |  |
| before bed | 47.5 ± 162 | 3.91 ± 9.29 | 1.00 |
| upon wake | -5.35 ± 15.3 | 0.93 ± 11.7 | 0.482 |
| wake+30 min | 2.11 ± 16.9 | -4.86 ± 13.83 | 0.482 |

Values presented as mean ± SD. \* indicates $P \leq 0.05$ .

#### Sleep Regularity Index

For the trainees remaining in circadian alignment, Sleep Regularity Index was 65.5 ± 13.4 during in-class training and changed to 67.0 ± 10.2 during field-training. Whereas for the trainees who underwent circadian misalignment, Sleep Regularity Index was 64.5 ± 8.2 during in-class training and increased to 55.0 ± 9.8 during field-training. Circadian misalignment resulted in a significantly larger decrease in Sleep Regularity Index (*P*=0.050, Table 2)

### Salivary Assessments

Salivary data were analyzed from 19 participants. For the trainees remaining in circadian alignment, cortisol measured 30 minutes after waking was 0.31 ± 0.15 µg/dL during in-class training and changed to 0.35 ± 0.16 µg/dL during field-training. Whereas for the trainees who underwent circadian misalignment, cortisol measured 30 minutes after waking was 0.57 ± 0.25 during in-class training and decreased to 0.25 ± 0.14 µg/dL during field-training. Circadian misalignment resulted in a significantly larger decrease in cortisol measured 30 minutes after waking (*P*=0.0002, Table 2). Cortisol measures before sleep and upon waking did not significantly change during circadian alignment versus circadian misalignment (Table 2).

Uric acid measured before bed, upon waking, and 30 minutes after waking, testosterone measured before bed, upon waking, and 30 minutes after waking, and melatonin measured before bed, upon waking, and 30 minutes after waking did not significantly change during circadian alignment versus circadian misalignment (Table 2).

## DISCUSSION

Night shift work is associated with increased chronic disease risk (Antunes *et al*., 2010; Pan *et al*., 2011; Lieu *et al*., 2012; Barbadoro *et al*., 2013; Depner *et al*., 2014; Vetter *et al*., 2016; Manohar *et al*., 2017; Shan *et al*., 2018; Gao *et al*., 2019; Dutheil *et al*., 2020; Rivera *et al*., 2020; Schilperoort *et al*., 2020; Maidstone *et al*., 2021). To further our understanding of the health risks associated with this highly prevalent occupational demand, we are in need of field-based methodologies that quantify behavioral circadian disruption (Depner *et al*., 2020). Here, we used a commercially available wrist-worn activity tracker to assess alterations in activity and sleep patterns occurring as a result of changing from a dayshift to a nightshift work schedule.

We developed a novel method to detect periods of sleep and wake. Using a data post-processing step, we are now able to use commercially available devices to assess behavioral patterns in shift workers. Consumer activity trackers have long been used in research settings to assess behavior in various patient populations (Adams *et al*., 2021; Bayoumy *et al*., 2021). However, this technology relies on external clock time and self-reported sleep time of the user to detect sleep and wake episodes. This approach has a risk of bias towards mislabeling periods of low activity during nighttime hours as “sleep”, which may not necessarily be accurate during nighttime shiftwork. Reciprocally, there is a risk of bias towards mislabeling actual sleep episodes as “wake”, when sleep occurs during daytime hours. Hence, we adapted this novel sleep labeling method to overcome these limitations. Our method relies on heart rate data as input information, which is a physiological indicator of activity, rather than anticipated bedtime, to label sleep/wake episodes. We had approximately 7,340 event epochs per person and 76.5 days of continuous data collected for each person. This high frequency of data points yields a more accurate prediction compared to input variables collected with low frequencies (Dunn *et al*., 2021). This approach is particularly advantageous for smaller samples sizes and was effective in our 36-person current sample size.

Behavioral regularity contributes to internal circadian timing, while behavioral irregularities contribute to mistiming, or internal circadian dyssynchrony (Bass & Lazar, 2016). Sleep and wake patterns, in addition to eating and exercise, are relevant behaviors impacting circadian timing (Bass & Lazar, 2016; Zhang *et al*., 2021). The Sleep Regularity Index was established as a tool to quantify the degree of sleep regularity in a group of older adults (mean age=68.7 ± 9.2 y) (Lunsford-Avery *et al*., 2018). The initial validation study reports that greater sleep irregularity was associated with ten-year cardiovascular disease risk, as well as greater obesity, hypertension, fasting glucose, hemoglobin A1c, and diabetes status (Lunsford-Avery *et al*., 2018). In our study, we compared the sleep regularity index assessed during circadian misaligned and circadian aligned behavioral conditions. As expected, we observed a decline in sleep regularity during night shiftwork. This decline in the sleep regularity index occurred concurrently with changes in activity and sleep patterns assessed using the novel sleep labeling method. These complementary findings support the use of our sleep labelling method as a novel tool to assess changes in activity and sleep behavioral patterns imposed by a night shift schedule.

Circadian rhythms are intrinsic, self-sustaining patterns generated by internal molecular clocks residing in virtually all cells of the body (Takahashi *et al*., 2008). The gold-standard for assessing circadian rhythm in human is the constant routine protocol (Duffy & Dijk, 2002). Several hormones also display 24-hour oscillating rhythms. Alternatively, the secretion patterns of these hormones—including cortisol (Hofstra & de Weerd, 2008)—can be used to infer circadian phase. In this study, we assessed salivary cortisol using self-administered saliva detection kits. We observed that salivary cortisol decreased during circadian misalignment. This occurred in parallel with the decline in sleep regularity index as well as changes alternations in Garmin-reported activity and sleep. In addition to cortisol, we examined changes in salivary testosterone, uric acid, and melatonin. Testosterone and uric acid were unchanged during circadian misalignment. Testosterone was highly variable, in part because 33% of the cohort was female; thus, we did not detect significant changes. These data seem to suggest that the behavioral irregularities resulting from of a night shift schedule occurred without changes in endogenous circadian phase.

Effective strategies combating increased disease risk associated with shiftwork are needed (Schilperoort *et al*., 2020). However, understanding individual behavioral patterns and effects in real life settings will required field-tested methods. To do so, we partnered with local police trainees and leveraged their established training schedule. We controlled for the stress of transitioning from in-class training to field-training through comparisons of both nightshift and dayshift schedules. Aberrations observed in physiological parameters such as heart rate and cortisol can be used to evaluate the impact of circadian dyssynchrony on health parameters. Overall, there was high compliance to the study protocol. This may be in part due to the fact that research staff largely conducted recruitment, consenting, and data collection at the work site (local police academy) and electronically rather than requiring in-patient laboratory visits. We seek to overcome a critical methodological barrier by quantifying circadian rhythm disruption in field-based settings. And as with any field-based study, there were some challenges. The melatonin assay requires a relatively high sample volume (100 µL), whereas other biomarkers require lower volumes (salivary cortisol: 25 µL; salivary testosterone: 25 µL; salivary uric acid: 10 µL). We initially used the oral swab method of sample collection; however, this did not capture adequate volume resulting in missing values for melatonin. After 12 participants, we switched to the passive drool method in efforts to collect larger volumes; yet, this still resulted in inadequate volume. Future field-based studies aiming to assess salivary circadian biomarker may consider cortisol as a reliable parameter of circadian disruption or dyssynchrony.

Our long-term goal is to address this need by developing an index, or composite score, to quantify the impact of behavioral circadian disruption in humans. The current work is the first step towards this goal. We adapted commercially available wearable devices for use in the shiftwork setting by improving the accuracy of sleep labelling. Using heart rate and activity data as input rather than external clock time we were able to accurately identify sleep and activity episodes during both daytime and nighttime. In line with alterations in activity and sleep patterns, we also observed declines in the sleep regularity index and lower salivary cortisol—an endogenous marker of circadian phase. These concurrent observations serve as internal validation of the novel sleep labelling method used to analyze wearable data. We believe this progress in using a wearable to assess circadian-related metrics within the context of the shiftwork setting will allow us to conduct field studies of the effects of circadian misalignment on measures of human health. Ongoing, we intend to incorporate other behaviors that impact circadian rhythm, such as the timing of meals and exercise, into a composite score quantifying circadian rhythm disruption (Wolff & Esser, 2012; Sato *et al*., 2019; Gabel *et al*., 2021). We anticipate validating this score against transcriptional and metabolic markers of tissue circadian phase. Such a metric may eventually be used to guide the development of techniques mitigating the adverse health consequences associated with shiftwork. Long-term, we expect this work will lead to healthier shiftwork populations, reduced healthcare costs, and reduced employee turnover.

## Data Availability

Data will be made available by the authors upon reasonable request.

## Data Availability Statement

Data available upon reasonable request from the authors.

